# A homozygous p.(Arg371Ser) mutation in *FICD* de-regulates AMPylation of the human endoplasmic reticulum chaperone BiP causing infancy-onset diabetes and severe neurodevelopmental delay

**DOI:** 10.1101/2022.05.14.22275020

**Authors:** Luke A. Perera, Andrew T. Hattersley, Heather P. Harding, Matthew N. Wakeling, Sarah E. Flanagan, Ibrahim Moshina, Jamal Raza, Alice Gardham, David Ron, Elisa De Franco

## Abstract

Dysfunction of the endoplasmic reticulum (ER) in insulin-producing beta cells results in cell loss and diabetes mellitus. Here we report on 5 individuals from three different consanguineous families with infancy-onset diabetes mellitus and severe neurodevelopmental delay caused by a homozygous p.(Arg371Ser) mutation in *FICD*. The *FICD* gene encodes a bifunctional Fic domain-containing enzyme that regulates the ER Hsp70 chaperone, BiP, via catalysis of two antagonistic reactions: inhibitory AMPylation and stimulatory deAMPylation of BiP. Arg371 is a conserved residue in the Fic domain active site. The FICD^R371S^ mutation partially compromises BiP AMPylation in vitro but eliminates all detectable deAMPylation activity. Overexpression of FICD^R371S^ or knock-in of the mutation at the *FICD* locus of stressed CHO cells result in inappropriately elevated levels of AMPylated BiP. These findings, guided by human genetics, highlight the destructive consequences of de-regulated BiP AMPylation and raise the prospect of tuning FICD’s antagonistic activities towards therapeutic ends.

## INTRODUCTION

Translocation channels, chaperones, protein modifying enzymes, and membrane trafficking components all maintain protein folding homeostasis in the endoplasmic reticulum and contribute to the integrity of secretion in eukaryotes (reviewed in: Braakman and Hebert, 2013). Whilst components of this machinery are common to most cells, its dysfunction in animals often manifests prominently as failure of the insulin-producing pancreatic beta cells to keep up with the needs of the organism. Whilst the basis for this apparent hypersensitivity of beta cells to ER stress is incompletely understood, rare mutations in 10 genes encoding components of this general ER proteostasis network are known to cause monogenic forms of diabetes (reviewed in: Shrestha et al., 2021; Yong et al., 2021). Given the broad role of protein secretion in animal physiology, these monogenic diseases are enriched in extra-pancreatic manifestations, often resulting in syndromic forms of diabetes mellitus associated with developmental abnormalities (reviewed in: Sanchez Caballero et al., 2021).

The Hsp70 chaperone BiP is an essential cog of the ER proteostasis network (reviewed in: Pobre et al., 2019). BiP is encoded by an essential gene, *HSPA5*, whose inactivation leads to early embryonic lethality in mice (Luo et al., 2006) but loss-of function alleles in several BiP co-factors are associated with beta cell dysfunction (Danilova et al., 2019; Synofzik et al., 2014). Thus, beta cells are sensitive to disruption of BiP’s chaperone network.

Unique amongst Hsp70 chaperones, BiP is regulated by AMPylation, a reversible covalent modification by which an AMP moiety from ATP is transferred onto a protein’s hydroxylated side chain. AMPylation (also known as adenylylation) is extensively exploited by bacteria to regulate endogenous processes (Kingdon et al., 1967) or host proteins in the context of infection (reviewed in: Woolery et al., 2010). Animal genomes encode a single ortholog of the widely disseminated family of Fic domain proteins that carry out AMPylation in bacteria (Worby et al., 2009; Yarbrough et al., 2009). Animal FICD is an ER-localised type II transmembrane protein whose active site faces the lumen.

There, it single-handedly carries out two functionally antagonistic reactions: BiP AMPylation (Ham et al., 2014; Preissler et al., 2015b; Sanyal et al., 2015) and deAMPylation (Casey et al., 2017; Preissler et al., 2017a).

AMPylation, which occurs on a single residue, Thr518 (Preissler et al., 2015b), locks BiP in a conformation with low substrate affinity and impaired responsiveness to the stimulatory effect of J-domain co-factors, effectively neutralising the chaperone (Preissler et al., 2015b; Preissler et al., 2017b). This biochemical feature is consistent with earlier observations that levels of modified (AMPylated) BiP are regulated physiologically: decreasing with mounting ER stress and increasing with stress resolution (Hendershot et al., 1988; Laitusis et al., 1999). This balancing act is critically-dependent on FICD (Casey et al., 2017; Preissler et al., 2017a) and arises from fine tuning of the enzyme’s active site to reciprocally regulate its two antagonistic functions (Perera et al., 2021; Perera et al., 2019).

Despite advances in the biochemical analysis of BiP AMPylation, its consequences in terms of organismal physiology are less well understood. Photosensitivity has been reported in flies lacking *ficd* (Moehlman et al., 2018; Rahman et al., 2012), but in knockout mice the phenotype is limited to weak immunological and learning deficits (McCaul et al., 2021). Here, we identify a rare missense, recessive mutation within FICD’s active site that is present within 5 individuals diagnosed with infancy-onset diabetes mellitus and neurodevelopmental abnormalities. This finding provides a rare insight into the functional consequences of deregulated BiP AMPylation.

## RESULTS

### The homozygous FICD p.(Arg371Ser) mutation lead to infancy-onset diabetes with neurodevelopmental abnormalities

Two siblings, born to related parents, were referred to the Exeter genomics laboratory for genetic testing on suspicion of Wolcott Rallison syndrome, a disorder caused by recessive variants in the *EIF2AK3* gene that encodes PERK a regulator or ER proteostasis (Delepine et al., 2000). The proband (Figure 1a) was diagnosed with diabetes at the age of 23 weeks. He also had severe developmental delay and skeletal abnormalities were suspected when he was 5 years old. His older brother was similarly affected and died aged 7 (cause unknown). Analysis of all the known neonatal diabetes genes, including *EIF2AK3*, did not identify any likely causative variant.

**Figure 1:**
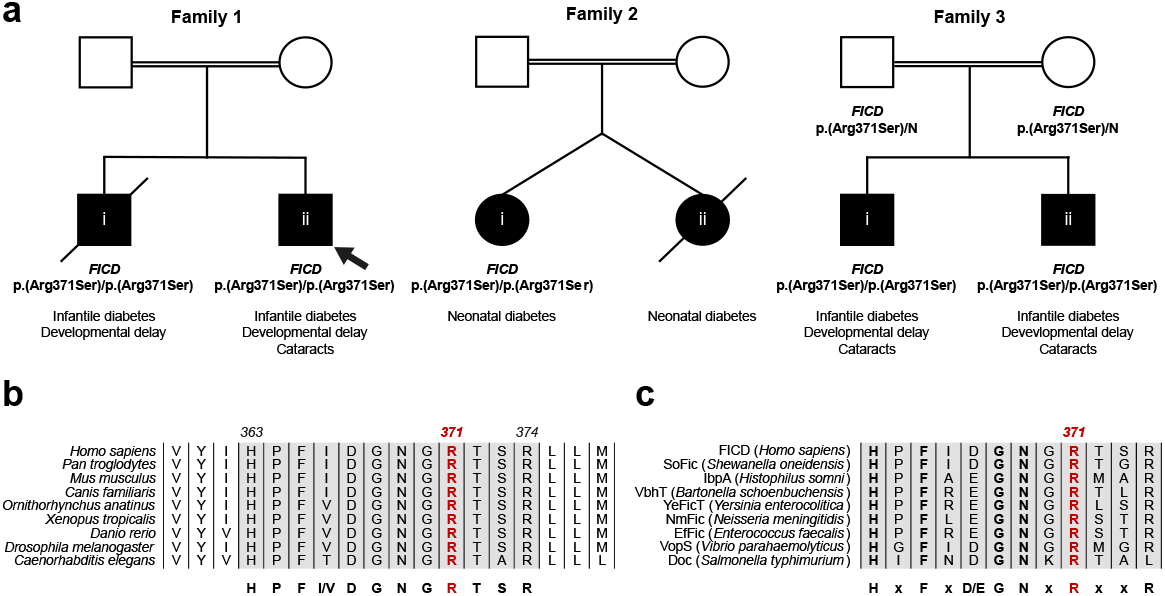
A homozygous p.(Arg371Ser) mutation within the human *FICD* gene results in infant-onset diabetes and severe neurodevelopmental delay. **a** Partial pedigrees of three families found to bear this rare genetic variant. Where available, information pertaining to *FICD* genotype and the phenotype of affected individuals are shown. The first individual identified to carry this homozygous mutation (the proband, family member 1ii) is indicated with an arrow. **b–c** Arg371 is located within the catalytic Fic motif and is highly conserved both across metazoan FICD orthologues **(b)**and across the wider Fic and Doc protein superfamily (**c**). The Fic motif is highlighted in grey, and residue numbering corresponds to human FICD.

Whole genome sequencing analysis of the two siblings was carried out to search for a novel genetic cause of monogenic diabetes. The two siblings were found to share 13 rare homozygous or hemizygous coding variants (Supplementary Table 1). None of these variants were located within genes previously linked to diabetes. We next investigated whether any of these 13 genes had been previously found to harbour homozygous or hemizygous variants in a cohort of 126 individuals diagnosed with diabetes before the age of 12 months, whose genome had been sequenced but no causative mutation discovered. This analysis revealed that one of the variants identified in the 2 siblings, the NM_007076.2:c.1113G>C, p.(Arg371Ser) in the *FICD* gene, had been previously detected in three additional individuals from 2 families, taking the total number of individuals homozygous for this variants to 5, all diagnosed with neonatal/infancy-onset diabetes (Figure 1a and Table 1).

**Table 1:** Summary of clinical features of the 5 individuals homozygous for the *FICD* p.(Arg371Ser) variant. The number of individuals for whom information on each feature was available is indicated (n).

| Cohort feature |  |
| --- | --- |
| Sex (n = 5) | Male (4) |
| Age at last assessment (mean, years) | 5.2 (range 0. – 9) |
| Birth weight (mean, g) (n=3) | 3063 (range 2800-3460) |
| Gestation (mean, weeks) (n=3) | 40 (range 38-41) |
| Infancy-onset diabetes (n=5) | 5 |
| Age at diabetes onset (mean, weeks) (n=4) | 29 (range 12-43) |
| Insulin dose (mean, U/kg/day) at last assessment (n=3) | 1.0 (range 0.7-1.2) |
| Developmental delay (n=4) | 4 |
| Cataracts (n=4) | 3 |
| Short stature (n=4) | 2 |

The p.(Arg371Ser) variant is very rare [minor allele frequency = 3.98 × 10^−6^ in the GnomADv2 database (Karczewski et al., 2020)] and it was not found in the homozygous state in either the ‘100,000 genomes’, ‘Gene and health’, ‘GnomAD’, ‘BioBank’, or ‘Decipher’ data sets. The p.(Arg371Ser) variant affects a residue located within the catalytic Fic domain motif and as such it is highly conserved both across metazoan FICD protein homologues (Figure 1b) and across the wider Fic and Doc (Fido) protein family (Figure 1c).

Since the 5 individuals were homozygous for the same *FICD* variant, we investigated whether they shared a haplotype, consistent with them being related and having inherited the variant from a common ancestor. Analysis of the whole genome sequencing data revealed that the four affected individuals in Family 1 and Family 3 shared a 4.5Mb haplotype that includes the *FICD* locus, suggesting that their *FICD* p.(Arg371Ser) variant was inherited from a common distant ancestor (Figure 1—figure supplement 1).

Within the haplotype, they shared three rare variants, of which *FICD* p.(Arg371Ser) was the only one predicted to affect the encoded protein. Importantly, the individual in Family 2 did not share a haplotype with the other two families, suggesting that the *FICD* variant is likely to have arisen independently in this family.

The clinical features of the 5 individuals homozygous for the *FICD* p.(Arg371Ser) mutation are summarised in Table 1. All cases were diagnosed with diabetes before the age of 12 months and needed replacement insulin therapy. The proband in Family 2 had a sibling who was also diagnosed with diabetes but died in the neonatal period (no DNA was available). The birth weight in this cohort was within the normal range (average 3,063g, range 2,800g-3,460g), consistent with these patients not having a severe reduction in fetal insulin secretion in utero. None of the parents in the 3 families (who are obligate heterozygous carriers of the *FICD* mutation) had diabetes.

Severe developmental delay was diagnosed in all the affected children except for one (the proband in Family 2) who could not be re-assessed after insulin-dependent diabetes was discovered at infancy. 3/5 affected children developed bilateral cataracts.

Additional features such as short stature, skeletal abnormalities, microcephaly, and deafness were also observed but these features were not seen in more than one family, therefore it is unclear whether they are features of the FICD-related disease.

The identification of the same rare variant in three families, across two separate haplotypes, with similar clinical features, supports the *FICD* p.(Arg371Ser) mutation being responsible for the observed phenotype.

### The Arg371Ser mutation compromises both enzymatic activities of FICD in vitro but leads to dominance of BiP AMPylation over deAMPylation

The Arg371 is an active site residue conserved not only in FICD, but also in the very large family of bacterial Fic-domain proteins and in the wider Fido superfamily (Figure 1b–c) (Khater and Mohanty, 2015; Kinch et al., 2009). In AMPylating FICD, Arg371 contributes to the binding of the β-phosphate of ATP, a co-substrate in the reaction, and presumably in AMPylation transition-state stabilisation (Figure 2a). In the deAMPylating enzyme, Arg371 engages the side chain of Glu234 in the conformation necessary for Glu234 to position a catalytic water molecule in-line for nucleophilic attack into the phosphodiester bond linking BiP(Thr518) and the AMP moiety (Figure 2b). Thus, the Arg371Ser mutation is poised to compromise both reactions.

**Figure 2:**
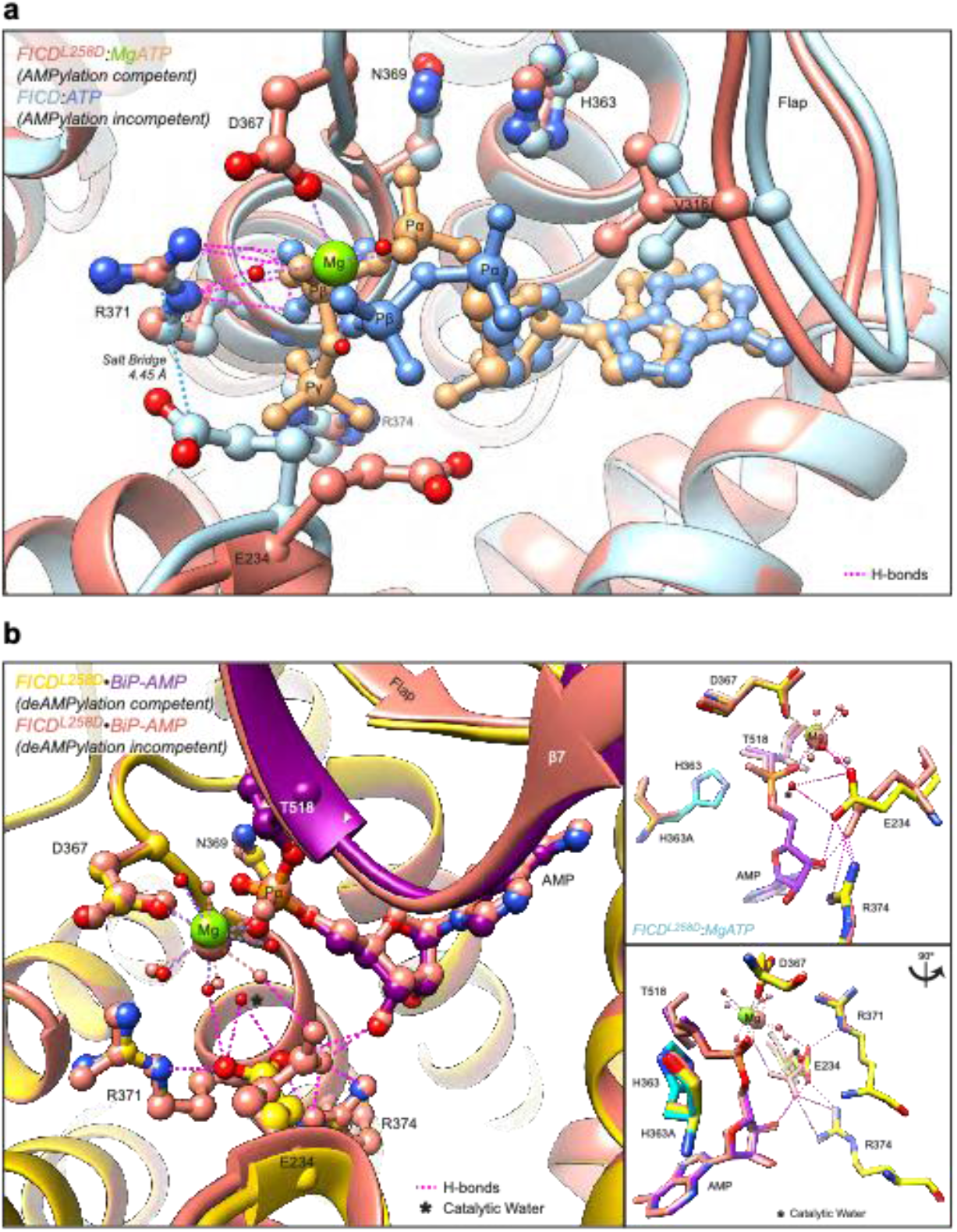
Mutation of Arg371 is predicted to perturb both FICD-catalysed AMPylation and deAMPylation. **a** Structures of monomeric (FICD^L258D^) and dimeric FICD proteins, crystallised in the presence of a large excess of magnesium cations and ATP, are superposed (PDB 6I7K and 6I7G, respectively). The former engages MgATP in an AMPylation competent conformation and the latter engages (only) ATP in an AMPylation incompetent conformation (Perera et al., 2019). Arg371 forms hydrogen bonds with both nucleotides (pink dashed lines). In addition, Arg371’s interaction with the β-phosphate of MgATP is also likely to help stabilise the AMPylation transition state. A potential long range ionic interaction between Arg371 and Glu234 in the non-competent conformation is also annotated. **b** Two alternative states of FICD’s active site engaged with its deAMPylation substrate, BiP-AMP. Glu234 can exist in at least two conformations. In the deAMPylation competent conformation, Glu234 correctly orientates a catalytic water molecule (*) for in-line nucleophilic attack into backside of the BiP-AMP phosphodiester bond (PDB 7B7Z). In the deAMPylation incompetent conformation Glu234 points away from the position of this putative catalytic water (PDB 7B80) (Perera et al., 2021). Hydrogen bonds formed by Glu234 are annotated in both cases (pink dashed lines). Arg371 interacts with Glu234 only when it resides in a deAMPylation competent conformation. Insets, reduced view of the superposed active sites, shown in orthogonal views, additionally overlaid with the catalytic His363 residues from PDB 6I7K (cyan).

The antagonistic nature of the two reactions implies that the phenotype arising from the mutation is likely to depend on the extent to which each is compromised. To address this issue, we expressed the globular luminal domains of wild-type and Arg371Ser mutant FICD in *E. coli* and evaluated the purified enzymes in vitro. FICD^R371S^ expressed well and demonstrated very similar elution profiles, as assessed by size exclusion chromatography, to the wild-type protein (Figure 3a). It was previously found that the oligomeric state of wild-type FICD reciprocally regulates its functionally opposed enzymatic activities (Perera et al., 2019). However, at the concentrations tested, both wild-type and mutant FICD appear principally dimeric, consistent with previous findings pertaining to wild-type FICD (Bunney et al., 2014; Perera et al., 2019). This suggests that differences in oligomeric state are unlikely to account for disparities between their relative enzymatic activities.

**Figure 3:**
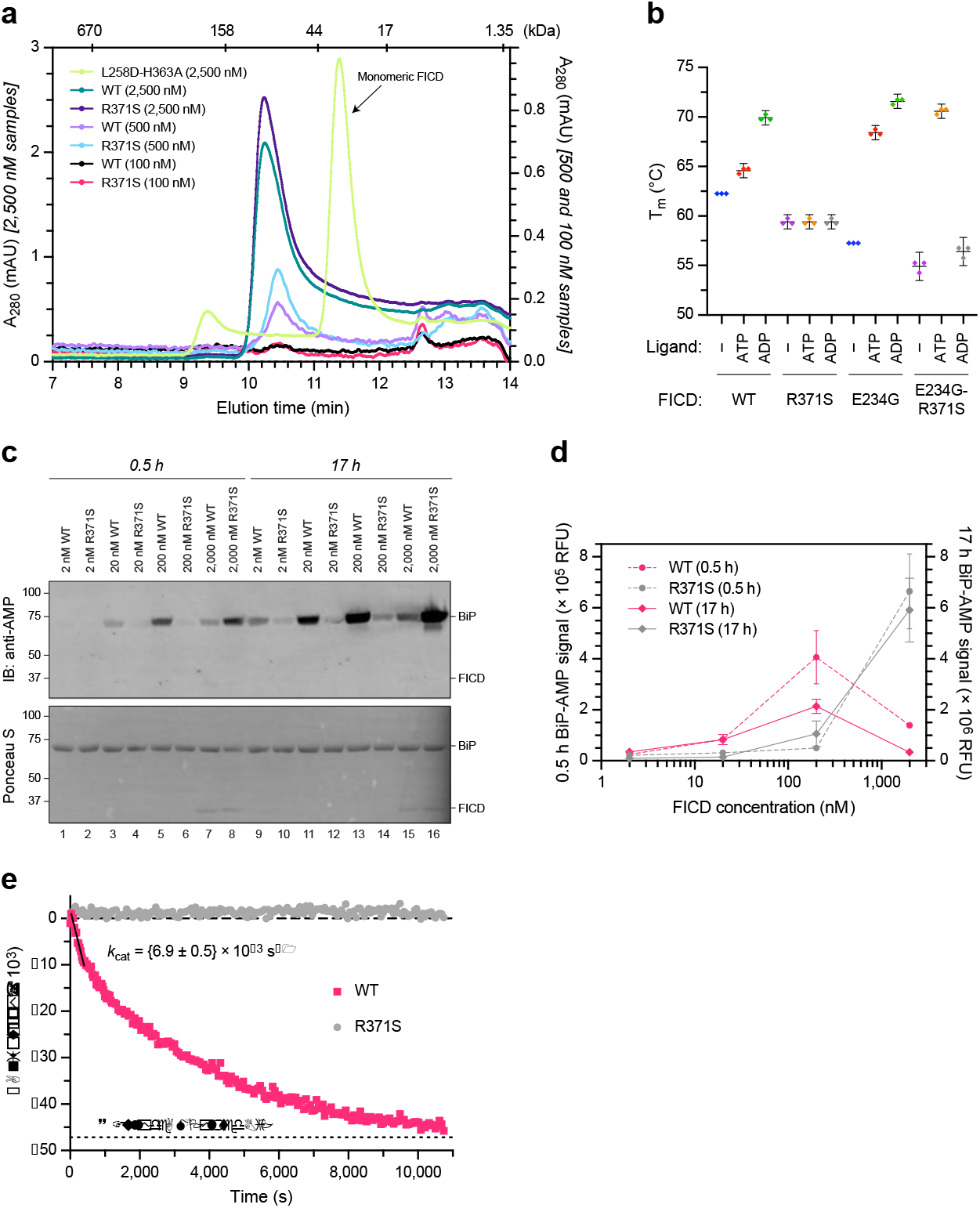
The FICD Arg371Ser mutation compromises BiP AMPylation and eliminates all detectable BiP deAMPylation in vitro. **a** Size exclusion chromatography confirms that the oligomerisation tendency of FICD^R371S^ is unperturbed relative to wild-type FICD. Across a range of assessable enzyme concentrations both FICDs elute as predominantly dimeric species. The elution profile of a monomeric FICD^L258D-H363A^ mutant is also shown for reference, alongside the elution times of molecular weight standards. **b** Plot of **t**he principal melting temperatures (T_m_s) of wild-type FICD (WT) and FICD^R371S^ in the presence or absence of nucleotide ligands and/or FICD’s gatekeeper Glu234 residue, derived from differential scanning fluorimetry (DSF). The mean T_m_ ± 95% confidence interval (CI) is displayed, from n = 3 independent experiments. Arg371Ser mutation slightly destabilises the apo enzyme and significantly reduces the ability of ATP and ADP to stabilise FICD. Glu234Gly mutation partially restores ATP (but not ADP) stabilisation of FICD^E234G-R371S^ (see figure supplement 1a). **c A r**epresentative immunoblot comparing the ability of wild-type and Arg371Ser FICD (at a wide range of enzyme concentrations) to catalyse the accumulation of BiP-AMP in the presence of physiological ATP concentrations (5 mM). At low enzyme concentrations (and early time points) the AMPylation defect imposed by Arg371Ser is manifest. At higher enzyme concentrations where the deAMPylation activity of the wild-type enzyme begins to dominate, the residual AMPylation activity of FICD^R371S^ results in a greater accumulation of BiP-AMP (relative to the same concentration of wild-type enzyme). **d** Quantification of the AMPylated BiP signals relative to total BiP (mean values ± SEM), from n = 4 independent experiments as represented in **c**. See figure supplement 1b. **e** A fluorescence polarisation-based deAMPylation assay highlighting the lack of activity catalysed by FICD^R371S^. Enzyme-saturating concentrations of BiP-AMP was provided as deAMPylation substrate and no enzymatic activity of 10 µM (shown) or 20 µM enzyme FICD^R371S^ was detectable. The estimated fluorescence anisotropy value of a fully deAMPylated BiP sample is derived from a heuristic fitting of a single exponential decay curve. The *k*cat value for wild-type FICD was calculated from n = 3 independent experiments (mean ± SEM).

Nucleotide free FICD^R371S^ possesses a modestly lower melting temperature than the wild-type enzyme, consistent with the stabilising effect of an ionic interaction between the side chains of Arg371 and Asp367 noted in the crystal structure of the apo wild-type enzyme (PDB 4U04).

Importantly, whereas the thermal stability of the wild-type enzyme increased significantly upon binding ATP and even more so ADP, the stability of the FICD^R371S^ mutant remained unchanged (Figure 3b and Figure 3—figure supplement 1). This finding is consistent with an important role for the Arg371 sidechain in contacting the β-phosphate of ADP or ATP bound in an AMPylation competent conformation or the γ-phosphate of ATP bound in an AMPylation incompetent conformation (Figure 2a) (Perera et al., 2019). Thus, the Arg371Ser mutation is likely to both lower the affinity of the FICD for nucleotide and reduce the protein-stabilising effect of any bound nucleotide di-or tri-phosphate.

The FICD^R371S^ mutant enzyme nonetheless retains some ability to bind nucleotide, as evidenced by the marked protein-stabilising effect of exogenous ATP when the Fic domain gatekeeper glutamate residue, that normally limits ATP affinity and inhibits AMPylation (Bunney et al., 2014; Engel et al., 2012), is also mutated (FICD^E234G-R371S^; Figure 3b and Figure 3—figure supplement 1a). Together these finding point to an important role of the Arg371Ser mutation in modifying the active site and leaves open the possibility of some residual enzymatic activityThe antagonistic activities of wild-type FICD are subordinate to its oligomeric state: the dimer is a good deAMPylase and a poor AMPylase and this trend is reversed in monomeric FICD (Perera et al., 2019). Given a *K*_d_ of FICD dimerisation in the nanomolar range, the relationship between the amount of AMPylated BiP and the FICD enzyme concentration, produced by reactions not otherwise limited by ATP, is biphasic — FICD’s AMPylation activity dominates at low (nanomolar) enzyme concentrations whilst its deAMPylation activity dominates at higher enzyme concentrations (Perera et al., 2019). This biphasic relationship is lost in reactions catalysed with FICD^R371S^. Moreover, at both early and late time points, more AMPylated BiP is recovered in reactions performed with the wild-type enzyme at low FICD concentrations, but this difference is reversed at higher FICD concentrations (Figure 3c–d and Figure 3—figure supplement 1b).

This situation is consistent with a defect in AMPylation activity of the mutant enzyme, which is perhaps most clearly demonstrated by the difference in BiP-AMP accumulation catalysed by low concentrations of FICD at early time points (conditions which are likely to report on the initial AMPylation rate of each enzyme). Nevertheless, the greater accumulation of AMPylated BiP catalysed by higher concentrations of FICD^R371S^, relative to FICD^WT^, is indicative of an even more significant impact of the mutation on FICD-mediated deAMPylation. Indeed, comparing the rate of BiP deAMPylation under substrate saturating conditions, by the wild-type and FICD^R371S^ enzyme, reveals robust activity of the former and no measurable deAMPylation activity of the latter (Figure 3e).

### FICD^R371S^ de-regulates BiP-AMP levels

Cells lacking FICD are unable to deAMPylate BiP (Casey et al., 2017; Preissler et al., 2017a). Therefore, the in vitro experiments above suggest that in cells expressing FICD^R371S^ as their only source of BiP-modifying enzymatic activity, the deAMPylation-unopposed, weak AMPylation function of the mutant FICD might induce a hyper-AMPylated pool of BiP. To test this idea, we transiently introduced wild-type and Arg371Ser mutant FICD into CHO cells lacking endogenous FICD and measured the effects on BiP AMPylation, using a recently described monoclonal antibody reactive with AMPylated proteins (MoAb 17G6, Hopfner et al., 2020).

Expression of wild-type FICD led to no detectable BiP-AMP signal, reflecting the dominance of its deAMPylating activity (Preissler et al., 2017a). By contrast the Arg371Ser mutant FICD promoted a conspicuous pool of AMPylated BiP (Figure 4a, compare lanes 2 & 4). The AMPylating activity of the Arg371Ser mutant was dependent on the integrity of its active site, as no BiP-AMP signal was detectable in cells expressing the double Arg371Ser-His363Ala mutant. As expected, levels of AMPylated BiP were even higher in cells expressing the deregulated Glu234Gly mutant FICD, which possesses both enhanced AMPylating activity and no detectable deAMPylating activity (Preissler et al., 2017a). The promiscuous features of FICD^E234G^ likely account for its auto-AMPylation and lower expression compared to the other FICD variants (Figure 4a, lane 3). The FICD immunoblot also reveals a minor, lower-mobility species in cells expressing FICD^R371S^ (Figure 4a, compare lane 2 with lanes 4 & 5, marked by an asterisk) likely reflecting cryptic glycosylation of Asn369, exposed by the mutation (see Discussion). Only the FICD^E234G^ cells manifested significant activation of their CHOP::GFP and XBP1::Turquoise unfolded protein response (UPR) reporters (Figure 4b), an observation consistent with a requirement for high levels of AMPylation to yield basal activation of the UPR (Perera et al., 2019).

**Figure 4:**
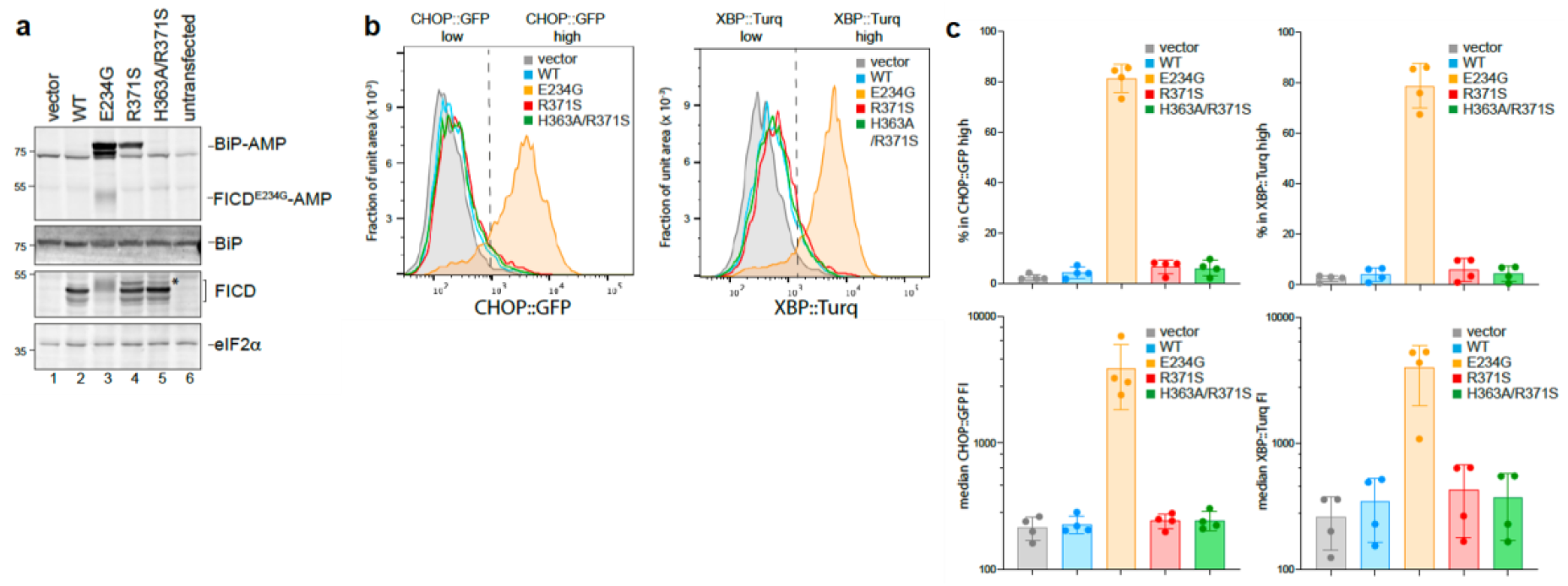
FICD^R371S^ promotes accumulation of AMPylated BiP in CHO cells lacking endogenous FICD. **a** Immunoblots of AMPylated proteins detected with a pan-AMP antibody, BiP, FICD and eIF2a (a loading control) in lysates of *FICD*^*Δ*^ CHO cells transiently transfected with plasmids encoding the indicated derivatives of FICD. Signals corresponding to AMPylated BiP, auto-AMPylated FICD^E234G^, total BiP, FICD and eIF2a are indicated. The novel species reactive with the anti-FICD antiserum in lysates of cells transfected with plasmids expressing FICD^R371S^ (horizontal arrow) likely reflects a glycosylated isoform arising from the creation of a new glycosylation site at Asn369. **b** Histograms of Integrated Stress response reporter CHOP::GFP and XBP::Turquoise (XBP::Turq, a reporter of the IRE1 branch of the UPR) fluorescence in *FICD*^*Δ*^ CHO dual reporter cells transfected with expression vectors as in “a.”. Transfected cells were gated for mCherry positivity, a marker carried on the expression plasmid in *trans* to the indicated FICD effector. **c** Quantification of the data from 4 repeats of the experiment shown in b. The upper two graphs show the percent of cells with high CHOP::GFP or high XBP::Turq for each transfection. The lower panel shows the mean +/-SD of the median fluorescence intensity for each reporter. (n=4)

To explore the consequences of more physiological expression levels of FICD^R371S^, we used CRISPR/Cas9 mediated homologous recombination to replace the endogenous wild-type FICD of CHO cells with an Arg371Ser mutant (Figure 5a).

**Figure 5:**
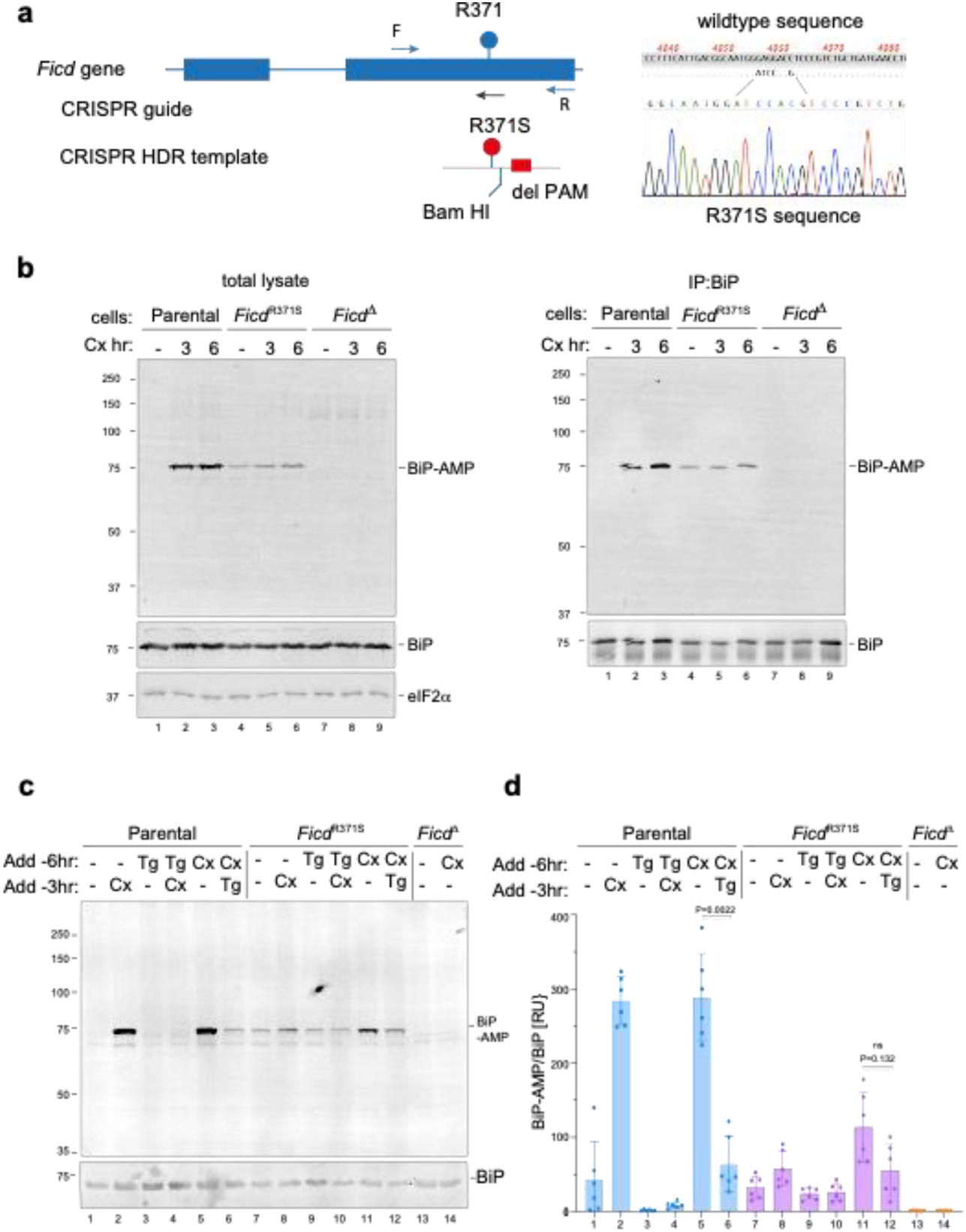
Expressed at the endogenous *FICD* locus, the p.(Arg371Ser) mutation leads to defective clearance of AMPylated BiP in ER stressed cells. **a** On the left is a schema of the Chinese Hamster Ovary cell *FICD* locus. Exons depicted as blue boxes, introns as lines. Forward (F) and reverse (R) primers used to genotype the locus and the guide RNA that directs the Cas9 nuclease (CRISPR guide) are depicted by arrows. A schema of the repair template is provided below, with the Arg371Ser mutation, the silent *Bam*HI site and silent PAM-destroying features noted. To the right is the sequence of the locus before and after recombination and a sequencing trace of the locus in the mutant *FICD*^*R371S*^ derivative cell line. **b** On the left are immunoblots of AMPylated proteins detected with a pan-AMP antibody, BiP and eIF2α (a loading control) in lysates of cells of the indicated genotype. The cells were untreated or exposed to the protein synthesis inhibitor, cycloheximide (Cx, 100 µg/mL) for the indicated time. In the panel on the right are immunoblots of samples immunoprecipitated with a BiP-specific antiserum from the lysates of the samples shown on the left. **c** Immunoblots as in “b” in lysates of cells of the indicated genotype. The cells were untreated or exposed to the protein synthesis inhibitor, cycloheximide (Cx, 100 µg/mL) and/or thapsigargin (Tg) for the indicated periods prior to harvest. **d** Quantification of ratio of the BiP-AMP to total BiP signal (expressed in arbitrary units) of repeats of the experiments in “c”. The ratio in each repeat, mean +/-SD of the BiP-AMP signal normalized to total BiP signal from the same samples, ‘P values by two tailed T test, n = 6).

The monoclonal antibody, reactive with AMPylated proteins, revealed a strong signal of 75 kDa in immunoblot of lysate from cycloheximide treated wild-type CHO cells and weaker signal in untreated cells (Figure 5b, left panel) a pattern consistent with previous observations (Laitusis et al., 1999; Preissler et al., 2015b). The identity of the signal with AMPylated BiP was confirmed by performing the same assay on BiP immunopurified from cells with a BiP-specific antibody (Figure 5b, right panel). *FICD*^*R371S*^ cells had a different profile of AMPylated BiP, with a baseline signal, that increased only modestly in cycloheximide-treated cells. Conversely, cells lacking FICD activity altogether (*FICD*^*Δ*^ cells) had only a background signal in the immunoblot, consistent with FICD’s essential role in BiP AMPylation (Preissler et al., 2015b).

To explore further consequences of the *FICD*^*R371S*^ mutation on the regulation of BiP AMPylation, we exposed wild-type and mutant cells to thapsigargin, an agent that induces ER stress by luminal calcium depletion. In wild-type cells the imposition of ER stress markedly lowered the BiP-AMP signal (compare lanes 1 with 3 and 5 with 6, Figure 5c) (as previously noted: Laitusis et al., 1999). By contrast, in *FICD*^*R371S*^ mutant cells the imposition of ER stress had a much weaker effect on BiP-AMP levels (compare lanes 7 with 9 and 11 with 12, Figure 5c–d). These observations are consistent with impaired ER stress-mediated de-AMPylation in the mutant cells and suggest the potential for defective proteostasis arising from their inability to adjust functional levels of BiP to fluctuating levels of ER stress.

## DISCUSSION

The findings presented here establish a specific recessive missense mutation [*FICD* p.(Arg371Ser)] in a gene encoding an ER-localised, protein modifying enzyme, as causing a previously unrecognised genetic syndrome characterised by infancy-onset diabetes mellitus and neurodevelopmental defects. The disease mechanism likely consists of an intra-organellar perturbation that compromises both insulin-producing beta cells and cells relevant to neurological development and/or function. Patients with pathogenic mutations in other genes known to cause diabetes through dysregulation of ER function also have early-onset insulin-requiring diabetes and neurodevelopmental features (Shrestha et al., 2021). The identification of the same mutation in three families suggests a mutation-specific mechanism, as opposed to overall loss of gene function. A biochemical analysis of the effects of the Arg371Ser mutation on FICD’s enzymology and the extant understanding of the ER chaperone BiP, a natural FICD substrate, suggest a mechanism for the underlying molecular pathology.

BiP, the only Hsp70 chaperone in the mammalian ER, is essential. Complete inactivation of the encoding gene compromises both unicellular eukaryotes such as budding yeast (Normington et al., 1989) and metazoans at an early developmental stage (Luo et al., 2006).

Reduction in BiP levels by exposure to SubA, a bacterial toxin that cleaves the protein intracellularly, is sufficient to compromise the secretory apparatus (Paton et al., 2006). BiP AMPylation on Thr518, locks the chaperone in an inert conformation that precludes productive engagement of substrate (Preissler et al., 2017b; Wieteska et al., 2017) and is thus tantamount to a reduction in the concentration of active BiP in the ER.

Normally, this process is subject to tight regulation — surplus BiP is inactivated by AMPylation as ER stress wanes and the reserve pool of AMPyated BiP is reactivated, through deAMPylation, as stress levels rise (reviewed in: Preissler and Ron, 2018). The Arg371Ser mutation compromises both aspects of BiP regulation, both timely BiP AMPylation and deAMPylation. The defect in AMPylation capacity explains the diminished ability of mutant cells to rapidly inactivate excess BiP upon a pharmacological reduction in unfolded protein flux into the ER (Figure 6).

**Figure 6:**
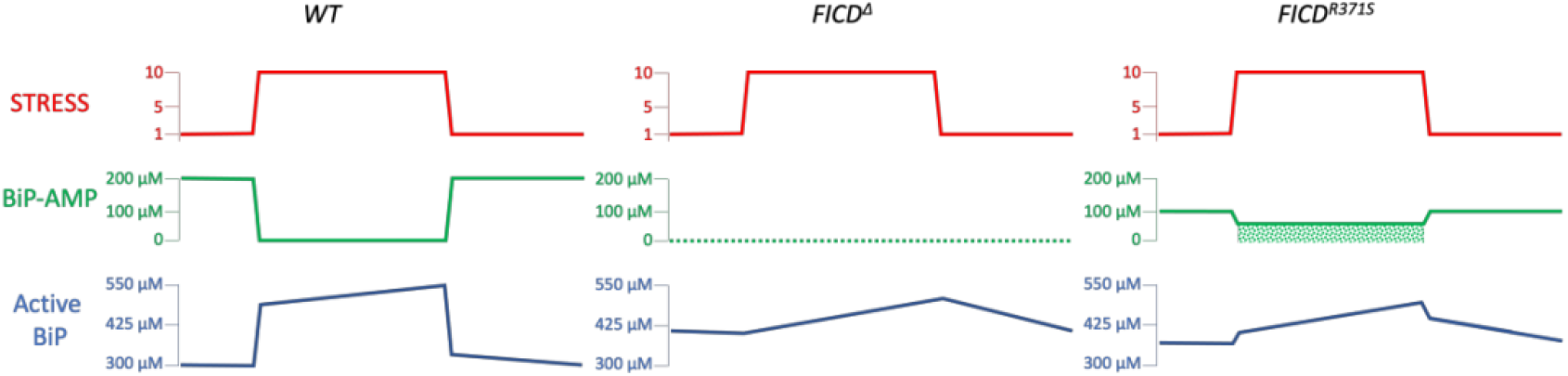
Cartoon contrasting the hypothetical divergent consequences of total absence of FICD (FICD^Δ^) with the Arg371Ser mutation (FICD^R371S^) in a pancreatic islet beta cell experiencing a physiologically-driven ten-fold increase in proinsulin biosynthesis (Itoh and Okamoto, 1980). In wild-type cells (*WT*) the stress imposed by increased production of proinsulin is met by rapid de-AMPylation of BiP and a slow, transcriptionally-mediated increase in BiP synthesis. As the stress wanes, BiP is re-AMPylated followed by slow decline in BiP protein levels, as its biosynthesis decreases. Note, the changes in concentration of BiP-AMP depicted here are based on measurements made in fed and fasted mice (Chambers et al., 2012) and assume a total concentration of BiP ∼500 µM. In *FICD*^*Δ*^ cells protein folding homeostasis is presumably maintained by transcriptional mechanisms that are adequate to sustain glycaemic control in knockout mice (McCaul et al., 2021). The *FICD*^*R371S*^ cells are burdened with a residual pool of AMPylated BiP, even when stressed (depicted by stippled green rectangle) compromising ER function.

The complete absence of deAMPylation activity of the mutant enzyme creates an intrinsic imbalance that favours AMPylation over deAMPylation to deprive cells of a readily accessible pool of dormant BiP when needed. Such deregulation is suggested by a consistent basal levels of AMPylated BiP in *FICD*^*R371S*^ cells, whereas the basal signal varies in wild-type cells varies. Importantly, under stress conditions wild-type cells are able to fully re-activate pre-existing pools of AMPylated chaperone, whilst the mutant cells are impaired in this recruitment mechanism (compare the effects of CHX treatment on wild-type and mutant cells in Figure 5).

This scenario is consistent with the recessive nature of the genetic disease. Whilst the imbalance in antagonistic enzymatic activities is intrinsic to the FICD^R371S^ mutant enzyme, the product of a wild-type *FICD* allele, a powerful deAMPylase (Preissler et al., 2017a), would have no difficultly in buffering the weak AMPylation bias imposed by a single allele of the mutant *FICD*. The buffering task of the wild-type protein in heterozygotes is likely favoured not only by the weak AMPylation activity of FICD^R371S^ but also by the fact that the mutation creates a cryptic glycosylation site in the enzyme’s active site. Glycosylation on Asn369 is likely to compromise FICD folding and reduce the burden of AMPylation even further in the heterozygous state. By contrast, in the homozygous state, the weak AMPylation activity of FICD^R371S^ molecules that escaped co-translation glycosylation is unopposed.

High levels of AMPylation can inactivate enough BiP to compromise ER function (Casey et al., 2017; Moehlman et al., 2018) and to trigger a measurable increase in signalling in the ER unfolded protein response (UPR) (Preissler et al., 2015b). By contrast, the activities of fluorescent UPR reporters are unaltered under basal conditions in FICD^R371S^-expressing cells. This likely reflects their adaptation to the mutant allele and suggests that the consequences of deregulated AMPylation are not manifest in the basal state but rather as cells experience fluctuations in the burden of ER client proteins. Insulin-producing beta cells are known to face dramatic (glucose-driven) excursions in the burden of unfolded pro-insulin entering their ER (>10 fold increase over 30 minutes, Itoh and Okamoto, 1980). A defect in recruiting AMPylated BiP to the chaperone cycle as beta cells face physiological glycaemic excursions, may account for their particular sensitivity to this mutation.

As mentioned above, the Arg371Ser mutation of FICD introduces a cryptic glycosylation site within the catalytic Fic domain (at Asn369). In the homozygous mutant the effect of possessing constitutively AMPylated BiP (with no recourse for re-activation/deAMPylation) may be further compounded by the constitutive production of a fraction of the expressed FICD which becomes Asn369-glycosylated and is thus unlikely to assume a properly folded state. Both BiP hyper-AMPylation and a resulting sensitisation to a potential FICD proteinopathy phenotype are more likely to occur in tissues with naturally higher levels of FICD expression, such as within the secretory tissues of the pancreas, pituitary and salivary glands (Wang et al., 2019).

It is interesting to contrast the consequences of complete loss of FICD function, which has no effect on glycaemic control in mice (McCaul et al., 2021), with the homozygous human p.(Arg371Ser) mutation. From a mechanistic perspective, this suggests that loss of AMPylation/deAMPylation as a post-translational strand of the UPR can be buffered by parallel regulatory processes, whilst inability to antagonise constitutive AMPylation is poorly tolerated (Figure 6). It also implies that the profound physiological perturbation arising from homozygosity of the *FICD*^*R371S*^ mutation could be reversed by complete inactivation of FICD, for example by a small molecule that blocks the enzyme’s adenosine-binding site.

Here, we interpret the consequences of the FICD^R371S^ mutation considering the inactivating features of BiP AMPylation: assuming that BiP-AMP is completely inert, and that the phenotype arises from a deficit of active chaperone. However, it remains possible that BiP-AMP retains important biochemical activities whose deregulation contributes to the phenotype via biochemical gain-of-function features. For example, it is possible that constitutive AMPylation of BiP may result in inappropriate sequestration of BiP co-chaperones (away from the active/unmodified pool of BiP).

Our focus on BiP is justified by the observation that it is the main/only protein to undergo detectable incorporation of AMP (Carlsson and Lazarides, 1983’ and 17G6 immunoblots here). However, it is noteworthy that the therapeutic strategy articulated above is relevant even if the pathogenic consequences of the mutation were to be realised via deregulated AMPylation of other, yet to be discovered, FICD substrates. Finally, it is intriguing to consider that the severe consequences of a genetic disease, which highlights the cost of an extreme imbalance in BiP AMPylation, may hint at the therapeutic utility of re-balancing the AMPylation/deAMPylation poise of wild-type FICD in other extreme circumstances.

## Supporting information

Key Resource Table

Plasmids used in this study

## Data Availability

All data produced in the present study are available upon reasonable request to the authors

## ACKNOWLEDGEMENTS

We thank Dorothea Hopfner and Aymelt Itzen (Hamburg-Eppendorf University) for the generous gift of the 17G6 monoclonal antibody to AMP. We also thank Dr Richard Caswell (Exeter Genomics laboratory) for helpful discussion regarding the mutation mechanism.

Supported by a Wellcome Trust Principal Research Fellowship to D.R. (Wellcome 200848/Z/16/Z). A Diabetes UK RD Lawrence fellowship to E.D.F. (Grant number 19_0005971), and a Wellcome Trust Senior Research Fellowship to S.E.F. (105636/Z/14/Z). M.N.W. is in receipt of an Independent Fellowship from the Exeter Diabetes Centre of Excellence funded by Research England’s Expanding Excellence in England (E3) fund.

For the purpose of open access, the authors have applied a Creative Commons Attribution (CC BY) licence to any Author Accepted Manuscript version arising from this submission.

## AUTHOR CONTRIBUTIONS

LAP: Designed, executed and interpreted the biochemical experiments. Assisted in design and interpretation of the cell-based experiments, co-wrote the manuscript

ATH: Oversaw design and interpretation of genetic and clinical data. Contributed to interpretation of genetic data and writing of the manuscript.

HPH: Designed, executed and interpreted the cell-based experiments. Edited the manuscript

MNW: Designed the pipeline and carried out the bioinformatic analysis of the whole genome data

SEF: Contributed to genetic and clinical data analysis

MI, JR and AG: Provided samples and clinical information

DR: Oversaw the design and interpretation of the biochemical and cell-based experiments. Co-wrote the first draft of the manuscript

EDF: Designed and analysed the genetic investigations. Identified the causative gene and mutation and led genetic follow up.

Collected clinical data. co-wrote the first draft of the manuscript.

## METHODS

### Supplementary Table 2 list the key resources and Supplementary Table 3 list the plasmids used in this study

#### Subjects

Individuals with diabetes diagnosed before the age of 12 months were recruited by their clinicians for molecular genetic analysis in the Exeter Molecular Genetics Laboratory. The study was conducted in accordance with the Declaration of Helsinki, and all subjects or their parents/guardians gave informed consent for genetic testing.

#### Genetic analysis

Genome sequencing was performed on DNA extracted from peripheral blood leukocytes. The samples from individuals 1a and 1b were sequenced on an Illumina HiSeq X10 with a mean read depth of 41.2 and 38.3, respectively. The samples of 126 individuals were analysed by whole-genome sequencing on an Illumina HiSeq 2500 (n=8), HiSeq X10 (n=71) or with BGISeq-500 (n=47). The sequencing data were analysed using an approach based on the GATK best practice guidelines. GATK haplotypecaller was used to identify variants which were annotated using Alamut batch version 1.11 (Sophia Genetics) and variants which failed the QD2 VCF filter or had less than 5 reads supporting the variant allele were excluded. Copy number variants were called by SavvyCNV (Laver et al., 2022) which uses read depth to judge copy number states. SavvyVcfHomozygosity (https://github.com/rdemolgen/SavvySuite) was used to identify large (>3Mb) homozygous regions in the genome sequencing data. An in-house software was used to detect shared haplotypes (https://github.com/rdemolgen/SavvySuite).

#### Protein Purification

The structured regions of human FICD proteins (residues 104–445) and BiP^T229A-V461F^ (residues 27–635) were expressed in T7 Express *lysY/I*^*q*^ (NEB) *E. coli* cells as N-terminal His_6_-SUMO fusion proteins. The proteins were expressed and purified (Perera et al., 2021; Perera et al., 2019). In brief, the His6-SUMO tag was removed from the FICD/BiP fusion partner by Ulp1 cleavage following Ni-NTA affinity purification. The cleaved proteins of interest were further purified by anion exchange (using a RESOURCE Q 6 ml column [Cytivia]) and gel filtration (using a S200 Increase 10/300 GL column [Cytivia]). Proteins were concentrated in a final buffer consisting of 25 mM Tris pH 8.0, 150 mM NaCl and 1 mM TCEP (buffer TNT). The plasmids used to express the FICD and BiP proteins utilised in this study are detailed in Supplementary Table 3.

#### Analytical Size Exclusion Chromatography

Analytical size exclusion chromatography was conducted on an Agilent Bio SEC-3 HPLC column (300 Å pore size, 3 µm particle size, 4.6 × 300 mm) equilibrated in TNT buffer at 25 °C. The FICD proteins were diluted in TNT buffer to the indicated concentration (Figure 3a) and incubated for at least 1 h at room temperature before a 10 µl volume of the protein solution was injected (using an HPLC autosampler) onto the column. Protein was eluted at a flow rate of 0.3 ml/min.

#### Differential scanning fluorimetry (DSF)

DSF analyses were conducted (Perera et al., 2021; Perera et al., 2019) with minor modifications. Final DSF samples contained 1 µM FICD protein ± 2.5 mM nucleotide (as indicated in Figure 3b) in a buffer of HKM supplemented with 1.5 × SYPRO Orange protein gel stain (Thermo Fisher Scientific).

#### In vitro AMPylation assay

5 µM unmodified BiP substrate was incubated at 25 °C with FICD protein at the indicated concentration for the indicated time (Figure 3c) in a buffer consisting of HKM (25 mM HEPES-KOH pH 7.4, 150 mM KCl and 10 mM MgCl_2_) supplemented with 5 mM ATP. Reactions were quenched by the addition of LDS-PAGE sample buffer and heating to 70 °C. Samples containing 1 µg of BiP were loaded onto and resolved on a 4-12% SDS-PAGE gel and subsequently wet-transferred onto a PVDF membrane. Total protein was imaged using Ponceau S stain. The membrane was blocked with 1 × ROTI Block (Roth) diluted in water and then probed for 1 h at 25 °C with a mouse monoclonal IgG antibody reactive to AMPylated proteins (MoAb 17G6, Hopfner et al., 2020), diluted 1/1000 (*v/v*) in 1 × ROTI Block. The AMPylated BiP signal was imaged using an IRDye 800CW goat anti-mouse IgG secondary antibody [Li-Cor], diluted 1/2000 (*v/v*) in 1x ROTI-Block.

#### In vitro deAMPylation assay

The fluorescence anisotropy-based deAMPylation assay and the *k*cat calculation (for wild-type FICD) was conducted (Perera et al., 2021; Perera et al., 2019) with minor modifications. Namely, each deAMPylation reaction was carried out in a 15 µl volume containing 100 µM BiP^T229A-V461F^-AMP (a concentration previously found to be able to saturate wild-type FICD) and 10 nM BiP^T229A-V461F^ modified with FAM labelled AMP, in a buffer of HKM supplemented with 0.05% (*v/v*) Triton X-100. The assay was conducted in a 384-well non-binding, low volume, HiBase, black microplate (greiner bio-one). Wild-type and Arg371Ser FICD enzymes were added at t = 0 to a final concentration of 10 µM. Note, FICD^R371S^ enzyme at 20 µM was also found to not catalyse any discernible deAMPylation activity.

The fluorescence anisotropy of the FAM signal was recorded on a CLARIOstar Plus plate reader (BMG Labtech) exciting at 482-16 nm and top reading emission at 530-40 nm. A reference well containing only 10 nM N^6^-(6-Amino)hexyl-ATP-6-FAM (Jena Bioscience) was used to set the relative gains in the parallel and perpendicular emission channels (targeting 25 mP units). The deAMPylation time course was conducted with the CLARIOstar maintaining a temperature of 25 °C, whilst also mixing the sample plate using a 2 s double orbital shake after each kinetic cycle. The fluorescence anisotropy signal of fully deAMPylated BiP was estimated from the plateau value of a single exponential decay curve heuristically fitted to the deAMPylation trace catalysed by wild-type FICD.

#### Mammalian cell culture

The previously described cell lines CHO-K1, CHO-K1-FICD^D^ (Ham et al., 2014; Preissler et al., 2015b; Sanyal et al., 2015) and CHO-K1-FICD^R371S^ cells (described below and in Key Resource Table) were cultured in Nutrient mixture F-12 Ham (Sigma) supplemented with 10% (vol/vol) serum (FetalClone II; HyClone), 1 × Penicillin-Streptomycin (Sigma) and 2 mM L-glutamine (Sigma) at 37 °C and 5% CO2. The identity of the CHO cell lines have been authenticated using the criteria of A. successful targeting of essential genes using species specific CRISPR whole genome library, and B. sequencing of the wildtype or mutant alleles of the genes studied that confirmed the sequence reported for the corresponding genome. The cell lines tested negative for mycoplasma contamination using a commercial kit (MycoAlert (TM) Mycoplasma Detection Kit, Lonza). None of the cell lines is on the list of commonly misidentified cell lines maintained by the International Cell Line Authentication Committee.

#### FICD mutation knock in

Plasmid and oligonucleotide reagents referenced herein are listed in Supplementary file 1/plasmid list and Key Resources Tables respectively. A guide targeting *Ficd* was selected in the region of Exon 2(previously called exon 3) surrounding the codon for R371S into the CCTop -CRISPR/Cas9 target online predictor (Stemmer et al., 2015) and duplex DNA oligonucleotides (3035_g2_R371_S, 3036_g2_R371_AS) of the sequence was inserted into the pSpCas9(BB)-2A-mCherry_V2 plasmid (UK1610) to create guide plasmid UK2959. Cells in 80% confluent 6-well dishes were transfected 24 hours post plating with 0.5 μg guide plasmid (UK2959) and 1.5 μg of a single stranded oligonucleotide repair template (3037_FICD_R371S_repair_V2S) that contains base changes to introduce the R371S allele as well as silent mutations introducing a BamHI site and pam destroying mutation using Lipofectamine LTX (Invitrogen). Thirty-six hours post transfection the cells were washed with PBS, resuspended in PBS containing 4 mM EDTA and 0.5% (w/v) BSA, and mCherry-positive cells were individually sorted by fluorescence-activated cell sorting (FACS) into 96-well plates using a Melody Cell sorter (Beckman Coulter). Genomic DNA was isolated from 186 clones of which 171 produced a 296bp PCR product with primers 3003_cgFICD_F and 3004_cgFICD_R. Upon digestion with BamHI, 5 had the expected size fragments and sequencing of these identified a single clone (R371S-C74) with a single correct R371S mutant allele.

#### Cell treatment, transfection and analysis

Cells (80% confluent) were transfected 24 hours after plating with 2 μg of the indicated expression plasmids per well of a 6-well (35cm) dish using Lipofectamine LTX (Life Technologies) according to manufacturer’s instructions. The medium was exchanged after 24 hr and harvest for Flow cytometry or immunoblot were performed 36 hr after transfection (figure 4). For analysis of CHO wildtype, FICD^D^ or FICD^R371S^ cells, medium on 80% confluent 10cm dishes of cells was exchanged 1 hour prior to drug treatments of cells (100 μg/ml cycloheximide or 0.2 μM thapsigargin) of the indicated genotype for 3 or 6 hours prior to harvest as indicated in the figures.

For immunoblotting cells were washed twice in PBS-2 mM EDTA and collected in the same and then lysed as previously described (Preissler et al., 2015a) in 4 cell volumes HG buffer (50 mM HEPES–KOH pH 7.4, 150 mM NaCl, 2 mM MgCl2, 33 mM D-glucose, 10% (v/v) glycerol, 1% (v/v) Triton X-100 and protease inhibitors (2 mM phenylmethylsulphonyl fluoride (PMSF), 4 μg/ml pepstatin, 4 μg/ml leupeptin, 8 μg/ml aprotinin) with 100 U/ml hexokinase (from Saccharomyces cerevisiae Type F-300; Sigma). Lysates were cleared at 20,000g and 20 μg of protein was separated by PAGE and transferred to PDVF membranes. For Immunoprecipitation, 430 μg of lysate was precleared with 20 μl UltraLink Hydrazine Resin (Pierce cat. # 53,149) blocked with aniline for 1 hour at 4C followed by incubation with 20 μl UltraLink Hydrazine Resin on which BiP-specific chicken IgY antibodies have been covalently immobilized according to the 2015a). After washing three times with lysis buffer the proteins were eluted with laemmli buffer prior to PAGE and transfer to PDVF. The PVDF membranes were subsequently blocked in 1/10 dilution of ROTI®Block 10x, 5% BSA in TBS (50 mM Tris-Cl, pH 7.5) then sequentially incubated with primary antisera diluted in blocking buffer as follows: anti-AMP (MoAb 17G6) and eIF2α [mouse anti-eIF2α (Avezov et al., 2013)] hamster BiP [chicken anti-hamster BiP (Avezov et al., 2013)], and FICD [chicken anti-FICD (Ham et al., 2014; Preissler et al., 2015b; Sanyal et al., 2015)] were used at a dilutions of 1/1000, 1/5000 and 1/1000, and 1/1000 (v/v), respectively.

The membranes were then washed 3 times in TBS and incubated with secondary antisera linked to IR800 (1/2000) or Cy3 1/1000 in blocking buffer. The membranes were scanned with an Odyssey near-infra-red imager (LI-COR) to detect IR800 secondary antisera or Biorad ChemiDocTM MP Imaging system with v3.0.1.14 Image Lab Touch software to detect Cy3. Where applicable, IB band quantification was carried out using NIH-Image (Fiji) Gel tool. manufacturer’s instructions (Preissler et al.,

## List of supplementary materials

**Figure 1 – figure supplement 1.**
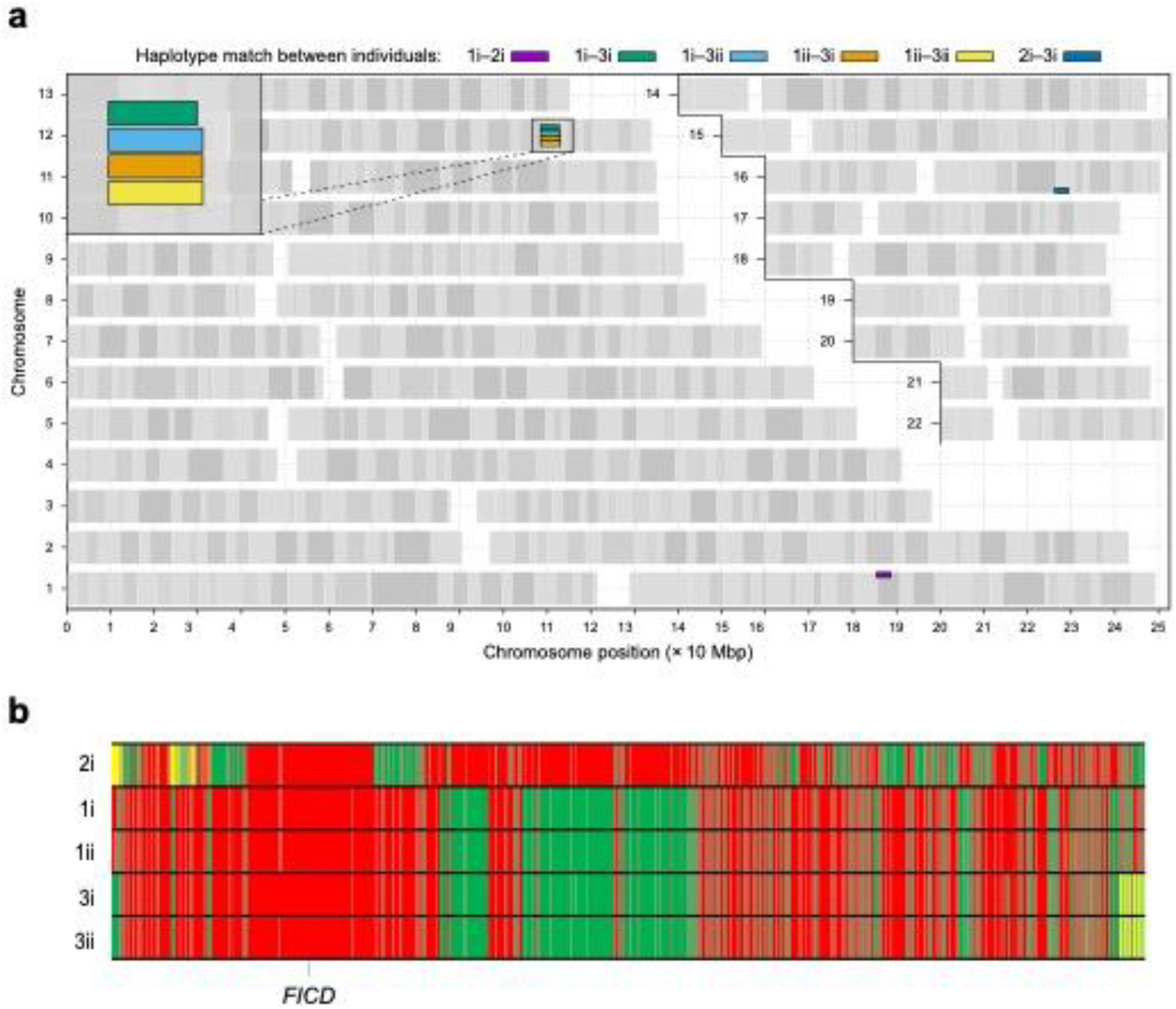
Analysis of shared haplotypes amongst affected individuals homozygous for the p.(Arg371Ser) mutation in *FICD*. **a** Graphical representation of genome-wide shared genomic segments larger than 3 Mbp in the 5 individuals with a *FICD* homozygous mutations. Each colour bar represents a haplotype shared by two individuals (across all 4 copies of the same chromosome), labelled according to Figure 1a. The 4 rectangles on the long arm of chromosome 12 (inset) show that the 4 individuals in Families 1 and 3 share a haplotype [approximate coordinates chr12(hg19):108,512,514–113,023,258] including the *FICD* gene [chr12(hg19):108,909,051–108,913,380], but none of them shares a haplotype with the patient in Family 2 (individual 2i) —hence the lack of annotated purple or dark blue bars. Note, genotype information was not available for family member 2ii. **b** Genotype calls for 2735 single nucleotide, not multiallelic variants, located on chr12:(hg19):108,500,000-113,100,000 in the 5 individuals with *FICD* homozygous p.(Arg371Ser) mutations. Only variants where at least one of the 5 patients carries the alternative allele are shown. Variants with a coverage of less than 15 reads and an allele balance for heterozygous calls <0.25 were removed. The position of the three variants in the *FICD* locus (including the pathogenic p.(Arg371Ser) variant) is indicated at the bottom of the graph. Green = homozygous for the reference allele, Yellow = heterozygous, Red = homozygous for the alternative allele. Note the similarity in variants pattern in affected individuals from families 1 & 3 and the dissimilarity with the affected member of family 2.

**Figure 3—figure supplement 1.**
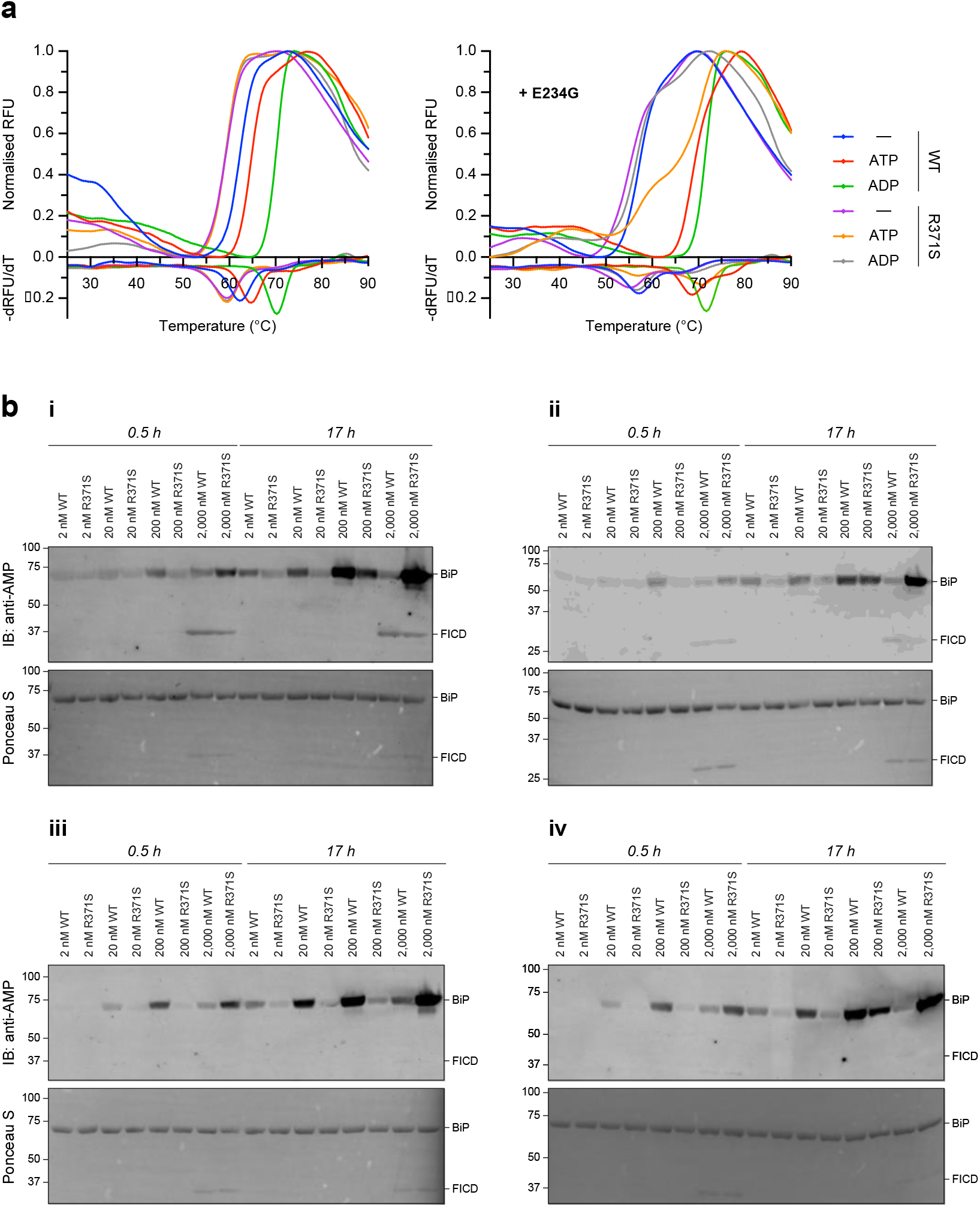
**a** Representative, normalised differential scanning fluorimetry (DSF) melt curves, shown above their corresponding negative first-derivatives. Note, FICD^E234G-R371S^ displays a non-uniform T_m_ shift in response to ATP. **b**, The 4 independent replicates of the immunoblot represented in **Figure 3c** (repeat **iii**) are shown. Note, the repeats (**i–iv**) are displayed in the order they were conducted.

## REFERENCES

Avezov, E., Cross, B.C., Kaminski Schierle, G.S., Winters, M., Harding, H.P., Melo, E.P., Kaminski, C.F., and Ron, D. (2013). Lifetime imaging of a fluorescent protein sensor reveals surprising stability of ER thiol redox. J Cell Biol 201, 337–349.

Braakman, I., and Hebert, D.N. (2013). Protein folding in the endoplasmic reticulum. Cold Spring Harbor perspectives in biology 5, a013201.

Bunney, T.D., Cole, A.R., Broncel, M., Esposito, D., Tate, E.W., and Katan, M. (2014). Crystal structure of the human, FIC-domain containing protein HYPE and implications for its functions. Structure 22, 1831–1843.

Carlsson, L., and Lazarides, E. (1983). ADP-ribosylation of the Mr 83,000 stress-inducible and glucose-regulated protein in avian and mammalian cells: modulation by heat shock and glucose starvation. Proceedings of the National Academy of Sciences of the United States of America 80, 4664–4668.

Casey, A.K., Moehlman, A.T., Zhang, J., Servage, K.A., Kramer, H., and Orth, K. (2017). Fic-mediated deAMPylation is not dependent on homo-dimerization and rescues toxic AMPylation in flies. J Biol Chem.

Chambers, J.E., Petrova, K., Tomba, G., Vendruscolo, M., and Ron, D. (2012). ADP ribosylation adapts an ER chaperone response to short-term fluctuations in unfolded protein load. J Cell Biol 198, 371–385.

Danilova, T., Belevich, I., Li, H., Palm, E., Jokitalo, E., Otonkoski, T., and Lindahl, M. (2019). MANF Is Required for the Postnatal Expansion and Maintenance of Pancreatic beta-Cell Mass in Mice. Diabetes 68, 66–80.

Delepine, M., Nicolino, M., Barrett, T., Golamaully, M., Lathrop, G.M., and Julier, C. (2000). EIF2AK3, encoding translation initiation factor 2-alpha kinase 3, is mutated in patients with Wolcott-Rallison syndrome. Nature Genetics 25, 406–409.

Engel, P., Goepfert, A., Stanger, F.V., Harms, A., Schmidt, A., Schirmer, T., and Dehio, C. (2012). Adenylylation control by intra-or intermolecular active-site obstruction in Fic proteins. Nature 482, 107–110.

Ham, H., Woolery, A.R., Tracy, C., Stenesen, D., Kramer, H., and Orth, K. (2014). Unfolded protein response-regulated Drosophila Fic (dFic) protein reversibly AMPylates BiP chaperone during endoplasmic reticulum homeostasis. J Biol Chem 289, 36059–36069.

Hendershot, L.M., Ting, J., and Lee, A.S. (1988). Identity of the immunoglobulin heavy-chain-binding protein with the 78,000-dalton glucose-regulated protein and the role of posttranslational modifications in its binding function. Mol Cell Biol 8, 4250–4256.

Hopfner, D., Fauser, J., Kaspers, M.S., Pett, C., Hedberg, C., and Itzen, A. (2020). Monoclonal Anti-AMP Antibodies Are Sensitive and Valuable Tools for Detecting Patterns of AMPylation. iScience 23, 101800.

Itoh, N., and Okamoto, H. (1980). Translational control of proinsulin synthesis by glucose. Nature 283, 100–102.

Karczewski, K.J., Francioli, L.C., Tiao, G., Cummings, B.B., Alfoldi, J., Wang, Q., Collins, R.L., Laricchia, K.M., Ganna, A., Birnbaum, D.P., et al. (2020). The mutational constraint spectrum quantified from variation in 141,456 humans. Nature 581, 434–443.

Khater, S., and Mohanty, D. (2015). In silico identification of AMPylating enzymes and study of their divergent evolution. Sci Rep 5, 10804.

Kinch, L.N., Yarbrough, M.L., Orth, K., and Grishin, N.V. (2009). Fido, a novel AMPylation domain common to fic, doc, and AvrB. PLoS One 4, e5818.

Kingdon, H.S., Shapiro, B.M., and Stadtman, E.R. (1967). Regulation of glutamine synthetase. 8. ATP: glutamine synthetase adenylyltransferase, an enzyme that catalyzes alterations in the regulatory properties of glutamine synthetase. Proc Natl Acad Sci U S A 58, 1703–1710.

Laitusis, A.L., Brostrom, M.A., and Brostrom, C.O. (1999). The dynamic role of GRP78/BiP in the coordination of mRNA translation with protein processing. J Biol Chem 274, 486–493.

Laver, T.W., De Franco, E., Johnson, M.B., Patel, K.A., Ellard, S., Weedon, M.N., Flanagan, S.E., and Wakeling, M.N. (2022). SavvyCNV: Genome-wide CNV calling from off-target reads. PLoS Comput Biol 18, e1009940.

Luo, S., Mao, C., Lee, B., and Lee, A.S. (2006). GRP78/BiP is required for cell proliferation and protecting the inner cell mass from apoptosis during early mouse embryonic development. Mol Cell Biol 26, 5688–5697.

McCaul, N., Porter, C.M., Becker, A., Tang, C.A., Wijne, C., Chatterjee, B., Bousbaine, D., Bilate, A., Hu, C.A., Ploegh, H., et al. (2021). Deletion of mFICD AMPylase alters cytokine secretion and affects visual short-term learning in vivo. J Biol Chem, 100991.

Moehlman, A.T., Casey, A.K., Servage, K., Orth, K., and Kramer, H. (2018). Adaptation to constant light requires Fic-mediated AMPylation of BiP to protect against reversible photoreceptor degeneration. eLife 7.

Normington, K., Kohno, K., Kozutsumi, Y., Gething, M.J., and Sambrook, J. (1989). S. cerevisiae encodes an essential protein homologous in sequence and function to mammalian BiP. Cell 57, 1223–1236.

Paton, A.W., Beddoe, T., Thorpe, C.M., Whisstock, J.C., Wilce, M.C., Rossjohn, J., Talbot, U.M., and Paton, J.C. (2006). AB5 subtilase cytotoxin inactivates the endoplasmic reticulum chaperone BiP. Nature 443, 548–552.

Perera, L.A., Preissler, S., Zaccai, N.R., Prevost, S., Devos, J.M., Haertlein, M., and Ron, D. (2021). Structures of a deAMPylation complex rationalise the switch between antagonistic catalytic activities of FICD. Nature communications 12, 5004.

Perera, L.A., Rato, C., Yan, Y., Neidhardt, L., McLaughlin, S.H., Read, R.J., Preissler, S., and Ron, D. (2019). An oligomeric state-dependent switch in the ER enzyme FICD regulates AMPylation and deAMPylation of BiP. EMBO J 38, e102177.

Pobre, K.F.R., Poet, G.J., and Hendershot, L.M. (2019). The endoplasmic reticulum (ER) chaperone BiP is a master regulator of ER functions: Getting by with a little help from ERdj friends. J Biol Chem 294, 2098–2108.

Preissler, S., Chambers, J.E., Crespillo-Casado, A., Avezov, E., Miranda, E., Perez, J., Hendershot, L.M., Harding, H.P., and Ron, D. (2015a). Physiological modulation of BiP activity by trans-protomer engagement of the interdomain linker. eLife 4, e08961.

Preissler, S., Rato, C., Chen, R., Antrobus, R., Ding, S., Fearnley, I.M., and Ron, D. (2015b). AMPylation matches BiP activity to client protein load in the endoplasmic reticulum. eLife 4, e12621.

Preissler, S., Rato, C., Perera, L.A., Saudek, V., and Ron, D. (2017a). FICD acts bifunctionally to AMPylate and de-AMPylate the endoplasmic reticulum chaperone BiP. Nat Struct Mol Biol 24, 23–29.

Preissler, S., Rohland, L., Yan, Y., Chen, R., Read, R.J., and Ron, D. (2017b). AMPylation targets the rate-limiting step of BiP’s ATPase cycle for its functional inactivation. eLife 6, e29428.

Preissler, S., and Ron, D. (2018). Early Events in the Endoplasmic Reticulum Unfolded Protein Response. Cold Spring Harbor perspectives in biology.

Rahman, M., Ham, H., Liu, X., Sugiura, Y., Orth, K., and Kramer, H. (2012). Visual neurotransmission in Drosophila requires expression of Fic in glial capitate projections. Nat Neurosci 15, 871–875.

Sanchez Caballero, L., Gorgogietas, V., Arroyo, M.N., and Igoillo-Esteve, M. (2021). Molecular mechanisms of beta-cell dysfunction and death in monogenic forms of diabetes. Int Rev Cell Mol Biol 359, 139–256.

Sanyal, A., Chen, A.J., Nakayasu, E.S., Lazar, C.S., Zbornik, E.A., Worby, C.A., Koller, A., and Mattoo, S. (2015). A novel link between Fic (filamentation induced by cAMP)-mediated adenylylation/AMPylation and the unfolded protein response. J Biol Chem 290, 8482–8499.

Shrestha, N., De Franco, E., Arvan, P., and Cnop, M. (2021). Pathological beta-Cell Endoplasmic Reticulum Stress in Type 2 Diabetes: Current Evidence. Front Endocrinol (Lausanne) 12, 650158.

Stemmer, M., Thumberger, T., Del Sol Keyer, M., Wittbrodt, J., and Mateo, J.L. (2015). CCTop: An Intuitive, Flexible and Reliable CRISPR/Cas9 Target Prediction Tool. PLoS One 10, e0124633.

Synofzik, M., Haack, T.B., Kopajtich, R., Gorza, M., Rapaport, D., Greiner, M., Schonfeld, C., Freiberg, C., Schorr, S., Holl, R.W., et al. (2014). Absence of BiP co-chaperone DNAJC3 causes diabetes mellitus and multisystemic neurodegeneration. Am J Hum Genet 95, 689–697.

Wang, D., Eraslan, B., Wieland, T., Hallstrom, B., Hopf, T., Zolg, D.P., Zecha, J., Asplund, A., Li, L.H., Meng, C., et al. (2019). A deep proteome and transcriptome abundance atlas of 29 healthy human tissues. Mol Syst Biol 15, e8503.

Wieteska, L., Shahidi, S., and Zhuravleva, A. (2017). Allosteric fine-tuning of the conformational equilibrium poises the chaperone BiP for post-translational regulation. eLife 6.

Woolery, A.R., Luong, P., Broberg, C.A., and Orth, K. (2010). AMPylation: Something Old is New Again. Front Microbiol 1, 113.

Worby, C.A., Mattoo, S., Kruger, R.P., Corbeil, L.B., Koller, A., Mendez, J.C., Zekarias, B., Lazar, C., and Dixon, J.E. (2009). The fic domain: regulation of cell signaling by adenylylation. Mol Cell 34, 93–103.

Yarbrough, M.L., Li, Y., Kinch, L.N., Grishin, N.V., Ball, H.L., and Orth, K. (2009). AMPylation of Rho GTPases by Vibrio VopS disrupts effector binding and downstream signaling. Science 323, 269–272.

Yong, J., Johnson, J.D., Arvan, P., Han, J., and Kaufman, R.J. (2021). Therapeutic opportunities for pancreatic beta-cell ER stress in diabetes mellitus. Nat Rev Endocrinol 17, 455–467.

