## Supplementary material for "A homozygous p.(Arg371Ser) mutation in *FICD* de-regulates AMPylation of the human endoplasmic reticulum chaperone BiP causing infancy-onset diabetes and severe neurodevelopmental delay": Supplememantal table 1

Supplemental table 1:

List of rare homozygous or hemizygous coding variants shared by the proband and his affected sibling

| Gene | Coding Effect | Allele depth in 1a | Allele depth in 1b | gNomenclature | cNomenclature | pNomenclature | GNOMAD MAF frequency | GNOMAD Hetz count | GNOMAD Homoz count |
|---|---|---|---|---|---|---|---|---|---|
| *ATXN3* | in-frame | 0,25 | 0,21 | Chr14(GRCh37):g.92537354_92537355insCTGCTGCTGCTGCTGCTGCTGCTGCTG | NM_001127696.1:c.870_871insCAGCAGCAGCAGCAGCAGCAGCAGCAG<br>NM_001127697.2:c.762_763insCAGCAGCAGCAGCAGCAGCAGCAGCAG<br>NM_001164774.1:c.232_233insCAGCAGCAGCAGCAGCAGCAGCAGCAG<br>NM_001164776.1:c.277_278insCAGCAGCAGCAGCAGCAGCAGCAGCAG<br>NM_001164777.1:c.112_113insCAGCAGCAGCAGCAGCAGCAGCAGCAG<br>NM_001164778.1:c.430_431insCAGCAGCAGCAGCAGCAGCAGCAGCAG<br>NM_001164779.1:c.552_553insCAGCAGCAGCAGCAGCAGCAGCAGCAG<br>NM_001164780.1:c.378_379insCAGCAGCAGCAGCAGCAGCAGCAGCAG<br>NM_001164781.1:c.705_706insCAGCAGCAGCAGCAGCAGCAGCAGCAG<br>NM_001164782.1:c.67_68insCAGCAGCAGCAGCAGCAGCAGCAGCAG<br>NM_004993.5:c.915_916insCAGCAGCAGCAGCAGCAGCAGCAGCAG | NM_001127696.1:p.Gln282_Gln290dup<br>NM_001127697.2:p.Gln246_Gln254dup<br>NM_001164774.1:p.Ala77_Gly78insAlaAlaAlaAlaAlaAlaAlaAlaAla<br>NM_001164776.1:p.Ala92_Gly93insAlaAlaAlaAlaAlaAlaAlaAlaAla<br>NM_001164777.1:p.Ala37_Gly38insAlaAlaAlaAlaAlaAlaAlaAla<br>NM_001164778.1:p.Ala143_Gly144insAlaAlaAlaAlaAlaAlaAlaAlaAla<br>NM_001164779.1:p.Gln176_Gln184dup<br>NM_001164780.1:p.Gln118_Gln126dup<br>NM_001164781.1:p.Gln227_Gln235dup<br>NM_001164782.1:p.Ala22_Gly23insAlaAlaAlaAlaAlaAlaAlaAlaAla<br>NM_004993.5:p.Gln297_Gln305dup<br>NM_030660.4:p.Gln242_Gln250dup | 0 | 0 | 0 |

| | | | | | | | | | |
|---|---|---|---|---|---|---|---|---|---|
| | | | | | NM_030660.4:c.750_751insCAGCAGCAGCAGCAGCAGCAGCAGCAG | | | | |
| *FADS6* | in-frame | 0,17 | 0,30 | Chr17(GRCh37):g.72889676_72889677ins72 | NM_178128.3:c.17_18ins72 | NM_178128.3:p.Pro15_Ala16insThrGluProMetGluProThrGluProMetGluProThrGluProMetGluProThrGluProMetGluPro | 0 | 0 | 0 |
| *NDUFAF8* | missense | 0,30 | 0,39 | Chr17(GRCh37):g.79214933T>G | NM_001353402.1:c.346T>G NM_001353403.1:c.184T>G | NM_001353402.1:p.Cys116Gly NM_001353403.1:p.Cys62Gly | 0 | 0 | 0 |
| *MADCAM1* | in-frame | 0,5 | 0,5 | Chr19(GRCh37):g.501785_501786insAGGAGCCTCCCGACACCACCTCCCAGGAGCCTCCCGACACCACCTCCC | NM_130760.2:c.784_785insAGGAGCCTCCCGACACCACCTCCCAGGAGCCTCCCGACACCACCTCCC | NM_130760.2:p.Ser261_Pro262insGlnGluProProAspThrThrSerGlnGluProProAspThrThrSer | 0 | 0 | 0 |
| *FICD* | missense | 0,38 | 0,37 | Chr12(GRCh37):g.108912988G>C | NM_007076.2:c.1113G>C | NM_007076.2:p.Arg371Ser | 3.98E-06 | 1 | 0 |
| *USH1G* | missense | 1,37 | 0,35 | Chr17(GRCh37):g.72916591C>T | NM_001282489.2:c.31G>A NM_173477.2:c.340G>A NM_173477.4:c.340G>A | NM_001282489.2:p.Val11Met NM_173477.2:p.Val114Met NM_173477.4:p.Val114Met | 1.59E-05 | 4 | 0 |
| *HEPH* | missense | 0,21 | 0,24 | ChrX(GRCh37):g.65475978C>T | NM_001130860.3:c.2711C>T NM_001282141.1:c.2135C>T NM_014799.3:c.1901C>T NM_138737.4:c.2864C>T | NM_001130860.3:p.Ala904Val NM_001282141.1:p.Ala712Val NM_014799.3:p.Ala634Val NM_138737.4:p.Ala955Val | 0 | 0 | 0 |
| *NAP1L3* | missense | 0,19 | 0,22 | ChrX(GRCh37):g.92927906T>G | NM_004538.5:c.398A>C | NM_004538.5:p.Glu133Ala | 4.92E-05 | 5 | 0 |
| *FGD6* | missense | 0,26 | 0,27 | Chr12(GRCh37):g.95566432C>G | NM_018351.3:c.2530G>C | NM_018351.3:p.Asp844His | 6.37E-05 | 16 | 0 |

| | | | | | | | | | |
|---|---|---|---|---|---|---|---|---|---|
| *CUX2* | missense | 0,32 | 0,22 | Chr12(GRCh37):g.111652019G>T | NM_015267.3:c.79G>T | NM_015267.3:p.Val27Phe | 3.19E-04 | 73 | 0 |
| *ZSWIM6* | in-frame | 0,20 | 0,6 | Chr5(GRCh37):g.60628618_60628623del | NM_020928.1:c.519_524del | NM_020928.1:p.Ala183_Ala184del | 4.77E-04 | 8 | 1 |
| *MAGEB16* | missense | 0,18 | 0,13 | ChrX(GRCh37):g.35820836C>T | NM_001099921.1:c.523C>T | NM_001099921.1:p.Pro175Ser | 5.84E-04 | 43 | 0 |
| *CDR1* | in-frame | 0,12 | 0,10 | ChrX(GRCh37):g.139865954_139865971del | NM_004065.2:c.561_578del | NM_004065.2:p.Trp191_Phe196del | 6.42E-04 | 72 | 0 |
