## Supplementary material for "A homozygous p.(Arg371Ser) mutation in *FICD* de-regulates AMPylation of the human endoplasmic reticulum chaperone BiP causing infancy-onset diabetes and severe neurodevelopmental delay": Key Resource Table

Table S2: Key Resource Table

| Reagent type (species) or resource | Designation | Source or reference | Identifiers | Additional information |
|---|---|---|---|---|
| Cell line (*Cricetulus griseus* ) | CHO-K1 | ATCC | RRID:CVCL_0214 | |
| Cell line (*Cricetulus griseus* ) | CHO-K1, FicdΔ CHOP::GFP, XBP:turquoise | PMID: 26673894 | S21-FICD -/- clone 10 | |
| Cell line (*Cricetulus griseus* ) | CHO-K1, *Ficd* R371S | This paper | Ficd-R371S-c74 | |
| Sequence-based reagent | 3037_FICD_R371S_repair_V2S | This paper | 3037 | TCAATTCAGAGGACGCCATGAACCTG CACCCAGTTGAGTTCGCAGCCTTAGC CCATTACAAACTTGTGTACATCCACCC TTTCATTGACGGCAATGGATCCACGT CCCGTCTGCTGATGAACCTGATCCTG ATGCAGGCAGGGTACCCCCCAATCAC CATACTCAAGGAGCAGAGGTCTGAG TACTACCATGTATT |
| Sequence-based reagent | 3003_cgFICD_F | This paper | 3003 | GGATGTCGAGAAGCAGATGCAGGA |
| Sequence-based reagent | 3004_cgFICD_R | This paper | 3004 | CCTCCGTACACTTGGCAATGAAGC |
| Sequence-based reagent | 3035_g2_R371_S | This paper | 3035 | CACCGCAGGTTCATCAGCAGACGGG |
| Sequence-based reagent | 3036_g2_R371_AS | This paper | 3036 | AAACCCCGTCTGCTGATGAACCTGC |
| Commercial assay or kit | MycoAlert (TM) Mycoplasma Detection Kit | Lonza | LT07-118 | |
| Chemical compound, drug | Thapsigargin | MERCK-milipore | 586005 | |
| Chemical compound, drug | Cycloheximide | MERCK-milipore | C7698 | |
| Chemical compound, drug | Hexokinase Type F-300 | Sigma | Cat. #: H4502 | |
| Commercial assay or kit | ROTI®Block 10x | Carl Roth | Cat. #: A151 | |
| Antibody | Mouse monoclonal anti-AMP (MoAb 17G6) | PMID: 33299971 | | WB (1:1000) |
| Antibody | Chicken polyclonal anti-FICD IgY | PMID: 26673894 | | WB (1:1000) |
| Antibody | Chicken polyclonal anti-BiP IgY | PMID:23589496 | | WB (1:1000) |
| Antibody | Donkey polyclonal anti-chicken IgY; IRDye 800CW Donkey anti-Chicken Secondary Antibody | Li-Cor | Cat. #: 926–32218 | WB (1:2000) |
| Antibody | Goat polyclonal anti-mouse IgG (H+L); IRDye 800CW Goat anti-Mouse IgG Secondary Antibody | Li-Cor | Cat. #: 926–32210 | WB (1:2000) |
| Antibody | Goat polyclonal anti-chicken IgY; Cy3-Donkey anti-Chicken Secondary Antibody | Jackson Immuno Research | Cat. #: 103-165-155 | WB (1:1000) |
| Software, algorithim | Prism 9 | GraphPad | | |
| Software, algorithim | SavvyCNV | https://github.com/rdemolgen/SavvySuite | | |
| Software, algorithim | SavvyVcfHomozygosity | https://github.com/rdemolgen/SavvySuite | | |
| Software, algorithim | GATK haplotypecaller | Alamut batch version 1.8 (Interactive Biosoftware, Rouen, France) | | |
