## Supplementary material for "A homozygous p.(Arg371Ser) mutation in *FICD* de-regulates AMPylation of the human endoplasmic reticulum chaperone BiP causing infancy-onset diabetes and severe neurodevelopmental delay": Plasmids used in this study

Table S3: L:ist of plasmids used in this study

| ID | Plasmid name | Description | Encoded protein | PMID |
|---|---|---|---|---|
| UK1314 | pCEFL_mCherry_3XFLAG_C | Mammalian expression of C-terminally 3XFLAG. Neomycin-resistance replaced by mCherry (under SV40 promoter control) | mCherry | 25858979 |
| UK1397 | hsHYPE_WT_pCEFL-mCherry | Mammalian expression of full-length human FICD (HYPE) in pCEFL marked with mCherry | FICD & mCherry | 26673894 |
| UK1398 | hsHYPE_E234G_pCEFL-mCherry | Mammalian expression of full-length human FICD (HYPE) with activating E234G mutation in pCEFL marked with mCherry | $FICD^{E234G}$ & mCherry | 26673894 |
| UK1479 | hsHYPE_45–458_E234G_pGV67 | Bacterial expression of N-terminally GST tagged hsFICD(45-458) for preparative scale AMPylation of BiP | $GST\text{-}FICD^{E234G}$ | 26673894 |
| UK1481 | hsHYPE_104–445_E234G_pSmt3_pET28b | Bacterial expression of N-terminally His6-Smt3 tagged hsFICD(104-445) with E234G mutation | $FICD^{E234G}$ | 31531998 |
| UK1610 | pSpCas9(BB)-2A-mCherry_V2 | CRISPR-Cas9- guide vector | Nuclease & guide | 29198525 |
| UK2052 | hsHYPE_104–445_pSmt3_pET28b | Bacterial expression of wild-type N-terminally His6-Smt3 tagged hsFICD(104-445) | FICD | 31531998 |
| UK2521 | haBiP_27–635_T229A_V461F_pSmt3_ pET28b | Bacterial expression of FL ATPase and substrate binding deficient His6-Smt3-BiP(27–635) | $BiP^{T229A\text{-}V461F}$ | 34408154 |
| UK2888 | hsHYPE_104–445_R371S_pSmt3_pET28b | Bacterial expression of N-terminally His6-Smt3 tagged hsFICD(104-445) with R371S mutation | $FICD^{R371S}$ | This study |
| UK2889 | hsHYPE_104–445_E234G_R371S_pSmt3_pET28b | Bacterial expression of N-terminally His6-Smt3 tagged hsFICD(104-445)_E234G with R371S mutation | $FICD^{E234G\text{-}371S}$ | This study |
| UK2959 | cgFICD_g2_R371_pSpCas9(BB)-2A-mCherry | CRISPR-Cas9- guide vector targeting CHO *Ficd.* Used for homology directed repair construction of R371S mutant | guide RNA | This study |
| UK2967 | hsHYPE_R371S_pCEFL | Mammalian expression of full-length human FICD (HYPE) with a R371S mutation in pCEFL marked with mCherry | $FICD^{R371S}$ & mCherry | This study |
| UK2968 | hsHYPE_H363A_R371S_pCEFL | Mammalian expression of full-length human FICD (HYPE) with R371S and H363A mutations in pCEFL marked with mCherry | $FICD^{H363A,\ R371S}$ & mCherry | This study |
